# Reported Drug Spectrum and Disproportionality Signals for Malignant Neoplasm Progression in FAERS: A Real-World Pharmacovigilance Study

**DOI:** 10.64898/2025.12.25.25343014

**Authors:** Yinghao Liu, Miao Zeng, Mingying Zhang, Hongxiang Xu, Xiaoyu Li, Jun Zhang

## Abstract

This study aimed to identify drugs disproportionately reported with malignant neoplasm progression, an uncommon but clinically important endpoint, using large spontaneous reporting systems. Public reports were analyzed from the FDA Adverse Event Reporting System (FAERS; 2004Q1–2024Q4) and the Japanese Adverse Drug Event Report database (JADER; 2004–2024). Cases were defined using MedDRA Preferred Terms for malignant neoplasm/tumour progression, and reports in which progression was recorded as an indication or medical history were excluded. Suspected drugs were standardized to generic names, and disproportional reporting was quantified using reporting odds ratios (RORs). Signals identified in FAERS were examined in JADER for cross-validation.FAERS contained 321, 020 progression-related reports, corresponding to 84, 977 unique cases after deduplication. Reporting increased over time and was associated with severe outcomes (death 27.63%; hospitalization 13.66%). Among the 50 drugs prioritized by report volume and signal strength, most were anticancer or immunomodulating agents (64%), and the highest report counts involved pembrolizumab, nivolumab, carboplatin, and enzalutamide. Disproportionality analysis detected positive signals for 41 drugs, with the strongest signals observed for afatinib, gefitinib, and osimertinib. In JADER (8, 929 cases), 22 of the 41 FAERS-positive signals were replicated with consistent direction but different magnitude.These findings are hypothesis-generating and suggest that tumor progression reporting clusters around specific therapies, particularly immunotherapies and targeted agents.These results support closer post marketing monitoring of selected drug event pairs and incentivize epidemiological and case-control studies to validate signals and elucidate clinical significance.

## 1 Introduction

Malignant tumor progression often determines the course of treatment and patient outcomes. Globally, cancer is a leading cause of death. In 2020, nearly 10 million deaths were attributed to cancer, accounting for about one in six deaths worldwide^[1]^. Even in high-income countries such as the United States, cancer remains the second leading cause of death after heart disease^[2]^.Treatment options have expanded in recent years. Immune checkpoint inhibitors (ICIs) are widely used in solid tumors and can improve survival in specific patients^[3]^. More than ten PD-1/PD-L1 or CTLA-4 antibodies have been approved worldwide for cancers such as lung cancer and melanoma^[4^]. Yet one question remains: does improved efficacy mean that the risk boundary is equally clear? Atypical outcomes have been observed. Some patients develop unexpectedly rapid tumor worsening after treatment, termed hyperprogressive disease (HPD)^[5]^. Hyperprogressive disease is often described as a sudden acceleration of tumor growth kinetics during immunotherapy, accompanied by rapid clinical decline. It is uncommon but clinically serious^[6]^. Systematic reviews report hyperprogressive disease rates of about 9.9%–18.06% across studies^[7]^. Patients with hyperprogressive disease have poorer outcomes than those with conventional progression, including shorter median overall survival^[7]^.The mechanism behind this type of atypical acceleration remains unclear, but there are still several factors have been linked to, including older age, liver metastases, elevated LDH, and low PD-L1 expression^[8][9]^. Baseline immune features and genomic alterations, such as specific T-cell phenotypes and MDM2/MDM4 amplification, have also been proposed^[10][11]^. For these reasons, abnormal progression has attracted increasing attention from both oncology and regulatory communities.

“Malignant neoplasm progression” is rarely listed as a typical adverse reaction in product labels. It is more often interpreted as lack of benefit or part of the natural course of cancer, rather than a direct toxic effect of a drug. Yet within the scope of this study, one question is hard to ignore: if progression is reported disproportionately often for a specific drug, should it not be treated as a potential safety clue^[12]^? Against this debated boundary of endpoint interpretation, large spontaneous reporting systems provide a useful, hypothesis-generating lens.

FAERS is among the largest spontaneous adverse event reporting systems worldwide, with more than ten million accumulated reports^[13]^. Submissions come from healthcare professionals, patients, and manufacturers, supporting post-marketing safety monitoring in real-world settings^[14]^. Compared with clinical trials, FAERS spans broader populations and longer time periods, which increases the chance of capturing rare, serious, or previously under-recognized events^[15]^. Prior FAERS-based analyses have reported potential signals for several drug classes, including proton pump inhibitors and targeted therapies^[16][17]^.

However, malignant neoplasm progression has seldom been treated as a primary endpoint in systematic mining. In many oncology safety analyses, progression or lack of benefit is excluded as a confounding outcome^[18]^.However, the excluded information may also contain clues indicating unusually rapid deterioration.To address this gap, we screened drugs reported with malignant neoplasm progression in FAERS, quantified disproportionality signals, and then examined the same signals in JADER to assess robustness. The goal was not to claim causality, but to identify drug–event pairs with concentrated progression reporting and to provide risk hypotheses for regulators and clinicians.

## 2 Materials and Methods

### 2.1 Data source and extraction

Within the scope of this study, we used publicly available spontaneous reports from FAERS (2004Q1–2024Q4) and the PMDA JADER database (2004–2024). FAERS data were processed according to FDA guidance. Quarterly files were downloaded, unpacked, and merged into a relational database with seven standard tables, including DEMO, DRUG, REAC, and OUTC^[19]^. Quarterly releases are not cumulative—what happens if they are simply summed across time? Version updates and duplicate submissions can be amplified, which can distort downstream signal estimates. To reduce this bias, reports were deduplicated by CASE_ID and only the latest case version was retained. Additional likely duplicates referring to the same event were removed using FDA-recommended procedures^[13][20]^. Deduplication reduces, but does not eliminate, residual duplication and missing fields; this constraint remains central when interpreting signals. After deduplication, malignant neoplasm progression reports in FAERS involved approximately 84, 977 unique cases.

### 2.2 Preferred Term definition

The endpoint was defined by MedDRA Preferred Terms (PTs): “Malignant neoplasm progression” and “Tumour progression.” REAC terms in FAERS are MedDRA-coded, so we identified reports containing either PT and extracted the corresponding cases. Records in which progression was coded as an indication or medical history were excluded to better align with adverse event reporting. This step also reflects a debated boundary: progression may represent an adverse outcome, but it can also reflect lack of benefit. In this study, progression was treated as the target adverse event for signal screening, without implying direct drug toxicity.

### 2.3 Drug definition

Suspected drugs were obtained from the DRUG table, including primary suspects (PS) and secondary suspects (SS). Because FAERS drug names are entered by both professionals and non-professionals, heterogeneous naming is common. Drug names were therefore standardized to generic names using the Medication Extraction System(MedEx). We summarized the frequency of each suspected drug in progression reports and used reporting volume and signal strength to prioritize drugs for focused review. The top 50 drugs were selected and grouped using the WHO Anatomical Therapeutic Chemical (ATC) classification to support interpretation of the therapeutic spectrum.

### 2.4 Disproportionality analysis

Disproportionality analysis are commonly used to screen for disproportionate reporting patterns in large pharmacovigilance databases^[21][22]^. We considered Proportional Reporting Ratio (PRR), Reporting Odds Ratio (ROR), Information Component (IC), and Empirical Bayes Geometric Mean (EBGM), and used ROR as the primary metric based on prior performance and interpretability in this setting^[23–25]^. Analyses were restricted to drug–event pairs with ≥3 reports. A positive signal was defined as a lower 95% confidence limit of the ROR >1 with a report count ≥3. Larger ROR values indicate stronger disproportionate reporting, not stronger proof of causality. For external cross-validation, the same endpoint definition, drug name normalization, and signal criteria were applied in JADER.

## 3 Results

### 3.1 Trend and gender-age analysis analysis

From Q1 2004 to Q4 2024, The FAERS database reports 321, 020 cases related to malignant neoplasm progression were identified, and 84, 977 unique cases remained after deduplication. Report counts increased over time, with a steeper rise after 2018 (Figure 1C). In spontaneous reporting systems, shifts in drug exposure, reporting intensity, and coding practices can occur together, so time trends are best treated as context rather than direct evidence of increased risk. One unexpected finding was that no entries for this endpoint were captured for 2011 in our extracted dataset.

**Fig 1.**
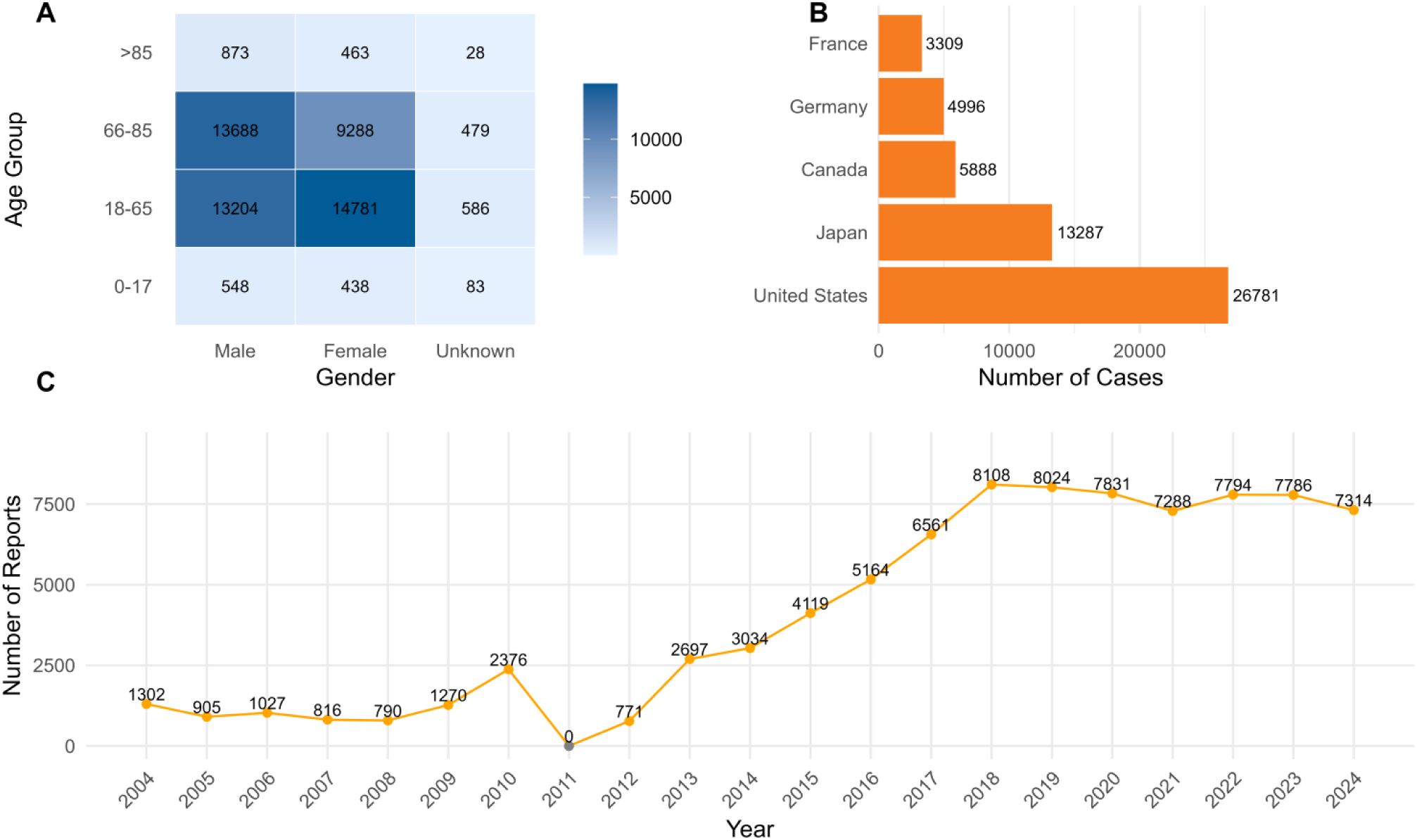
Clinical characteristics of reported cases of malignant tumor progression. (A) Heat map of Malignant Neoplastic Progression cases classified by age and gender; (B) Geographical distribution of Malignant Neoplastic Progression cases; (C) Annual Trends of Malignant Neoplastic Progression Report (2004-2024).

After excluding records with missing age or gender information, a total of 54, 459 cases were available for analysis. Most patients were 18–65 years (52.46%, n≈28, 571) or 66–85 years (43.07%, n≈23, 455) (Figure 1A). Patients aged ≤17 years and ≥86 years accounted for 1.96% and 2.50%, respectively. Females accounted for 45.85% (n≈24, 970) and males for 51.99% (n≈28, 313); sex was missing in 2.16%. The largest number of reports came from the United States (31.52%), followed by Japan (15.64%), Canada (6.93%), Germany (5.88%), and France (3.89%) (Figure 1B). Notably, serious outcomes were common, including death (27.63%) and hospitalization (13.66%). However, in severe clinical situations, disease progression is more likely to be reported, which should be taken into account when interpreting study results.

### 3.2 Drug distribution and therapeutic spectrum

Among the 50 prioritized drugs, antineoplastic and immunomodulating agents comprised 64% (32/50). Among the 32 anticancer-related drugs, targeted therapies were most common (43.8%), followed by cytotoxic chemotherapy (34.4%) and immunotherapies/other agents (21.9%) (Figure 2). The top ten reported suspected drugs were pembrolizumab, nivolumab, carboplatin, enzalutamide, dexamethasone, paclitaxel, everolimus, letrozole, oxycodone, and gemcitabine. Importantly, high report volume does not equate to high risk; it can also reflect high exposure and broad use across indications.

**Fig 2.**
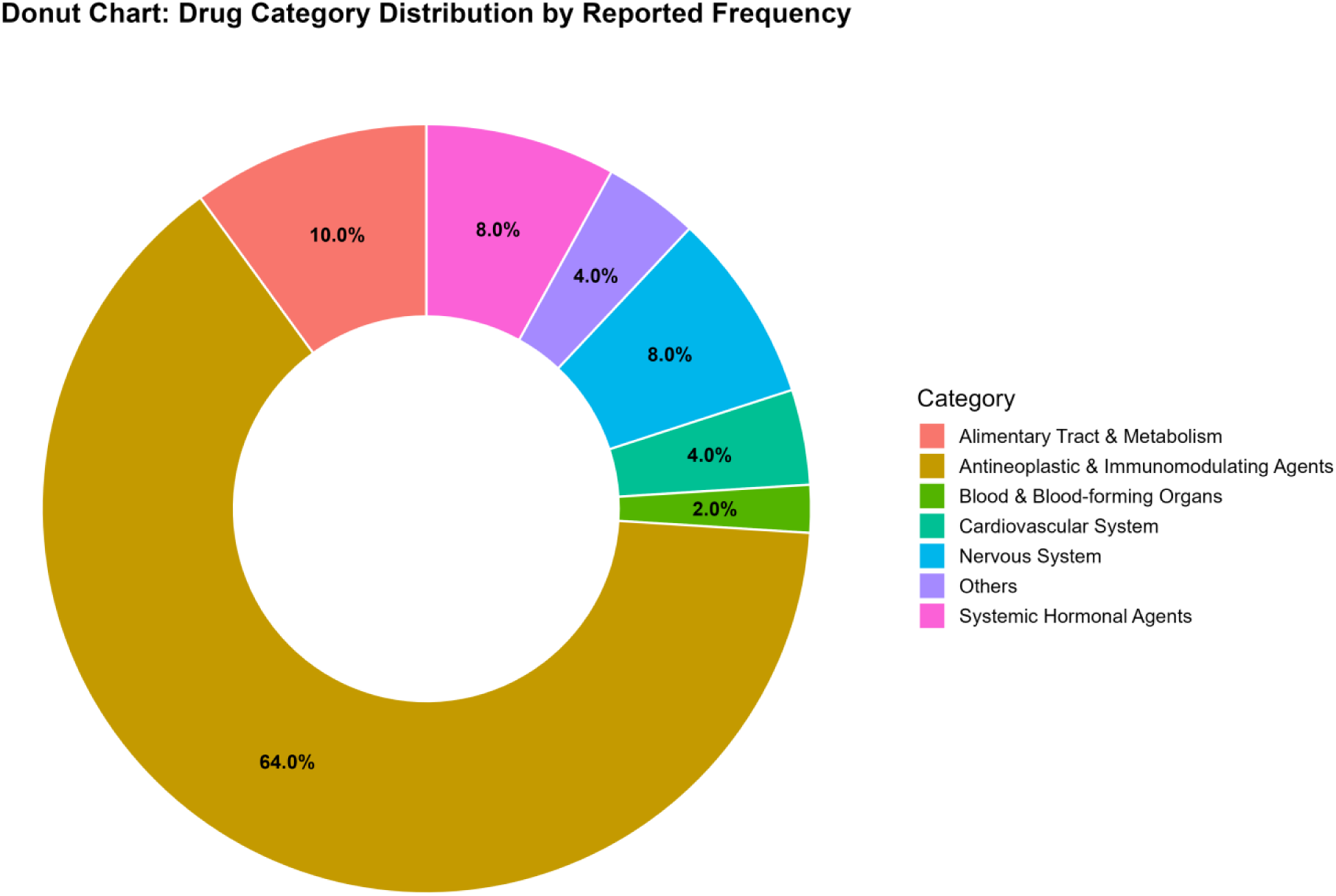
Classification of top 50 drugs associated with malignant tumor progression of nephrolithiasis.

By report count, the top 10 suspected drugs were pembrolizumab (n=9, 139), nivolumab (n=9, 068), carboplatin (n=5, 263), enzalutamide (n=5, 239), dexamethasone (n=4, 583), paclitaxel (n=4, 560), paracetamol (n=4, 539), everolimus (n=4, 495), letrozole (n=3, 886), and gemcitabine (n=3, 626). These agents are widely used in advanced cancers, including lung cancer, breast cancer, prostate cancer, renal cell carcinoma, melanoma, and Hodgkin lymphoma. At the class level, reporting was concentrated in platinum chemotherapy, taxanes, immune checkpoint inhibitors, endocrine therapies, and targeted therapies. This likely reflects high exposure and broad use of these drugs in standard and combination regimens, alongside the accumulation of malignant neoplasm progression reports.

### 3.3 Disproportionality analysis and external validation

To evaluate the potential association between drugs and malignant tumor progression, a disproportionality analysis was conducted on the top 50 drugs ranked by reporting frequency.ROR ranged from 0.55 to 50.32. Positive signals were observed for 41 drugs (≥3 reports and lower 95% CI of ROR >1). The strongest signals were for afatinib (ROR 50.32), gefitinib (ROR 45.56), and osimertinib (ROR 36.73) (Figure 3).

**Fig 3.**
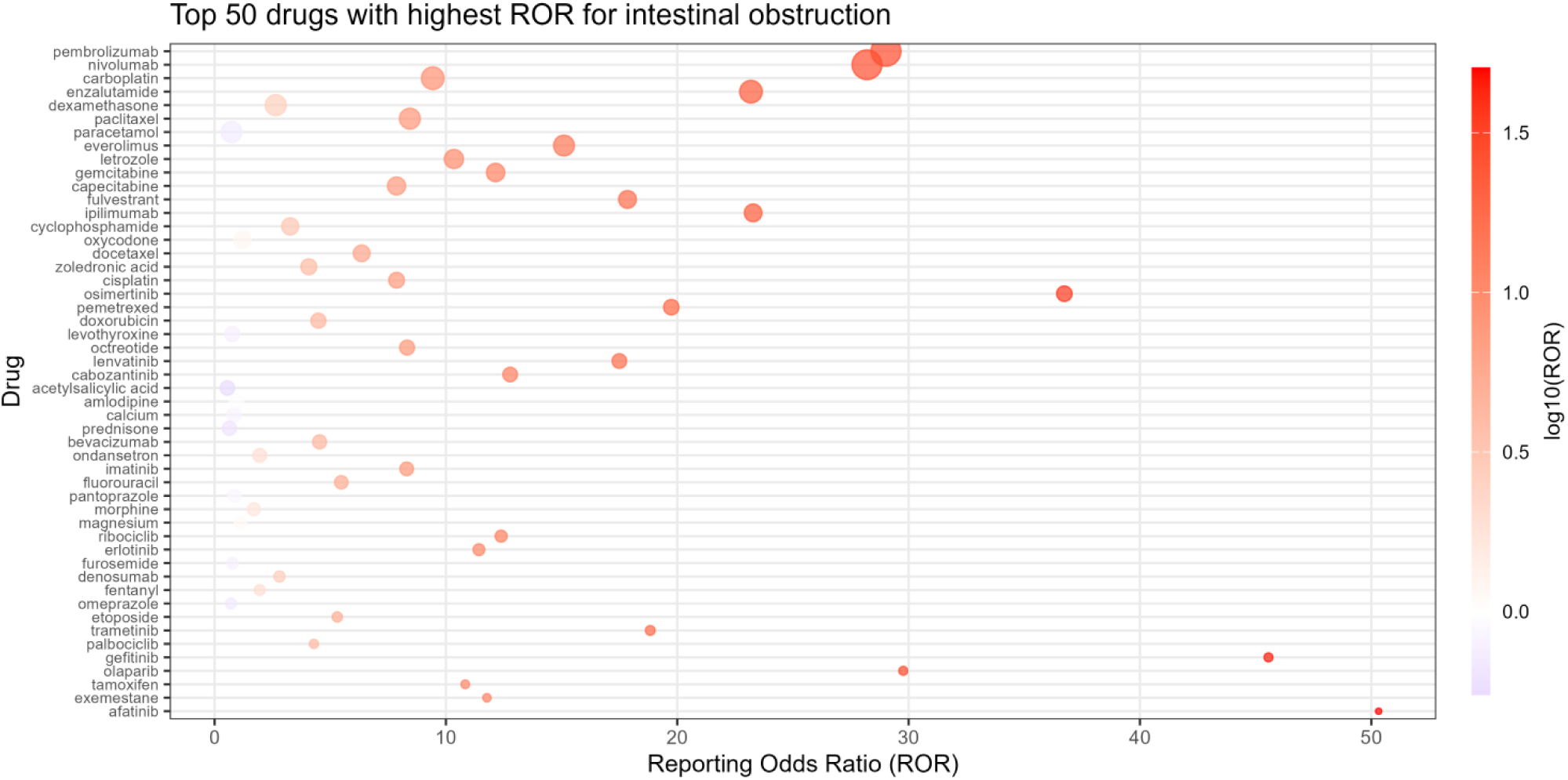
The distribution of ROR signal intensity associated with malignant tumor progression among the top 50 drugs.

In JADER, 8, 929 related cases were identified, and demographic patterns were broadly similar (Figure 4). Of the 41 FAERS-positive drugs, 22 were replicated in JADER with consistent direction but different magnitude (Figure 5). Because the prescribing structure, launch timing, indication mix, and reporting behavior can all shape signal strength; under the constraints of spontaneous reports, these differences are best viewed as robustness cues rather than direct biological evidence.

**Fig 4.**
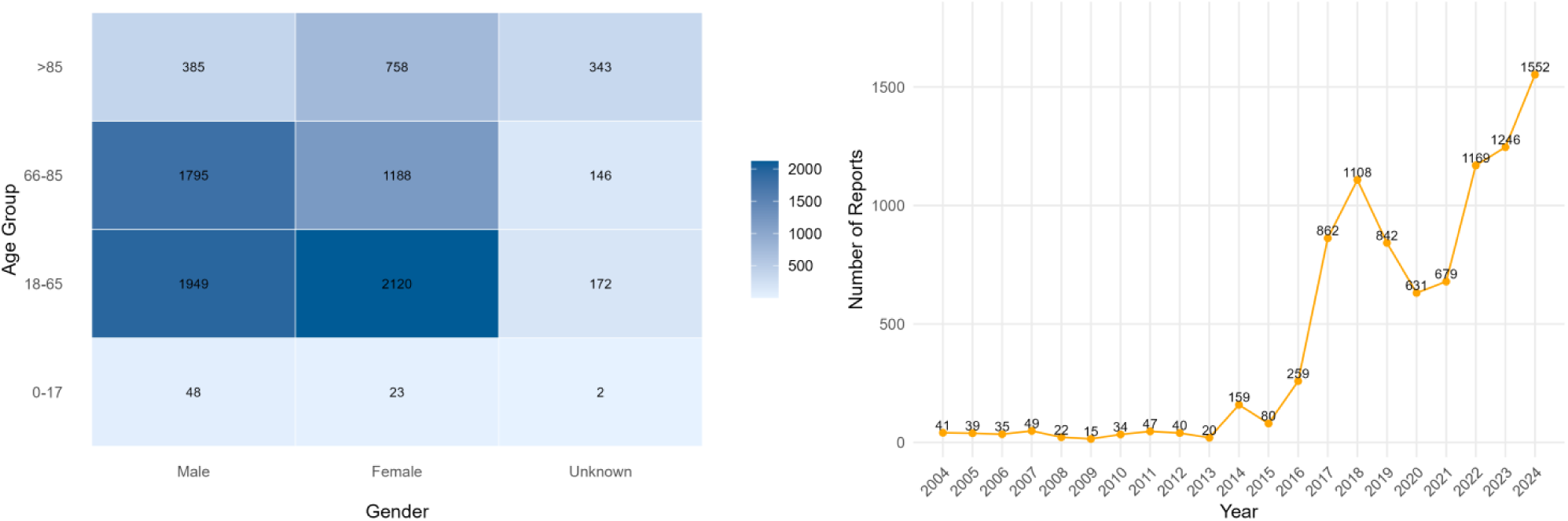
Clinical characteristics of reported cases of malignant tumor progression in Japan. (A) Heat map of Malignant Neoplastic Progression cases classified by age and gender; (B) Annual Trends of Malignant Neoplastic Progression Report (2004-2024).

**Fig 5.**
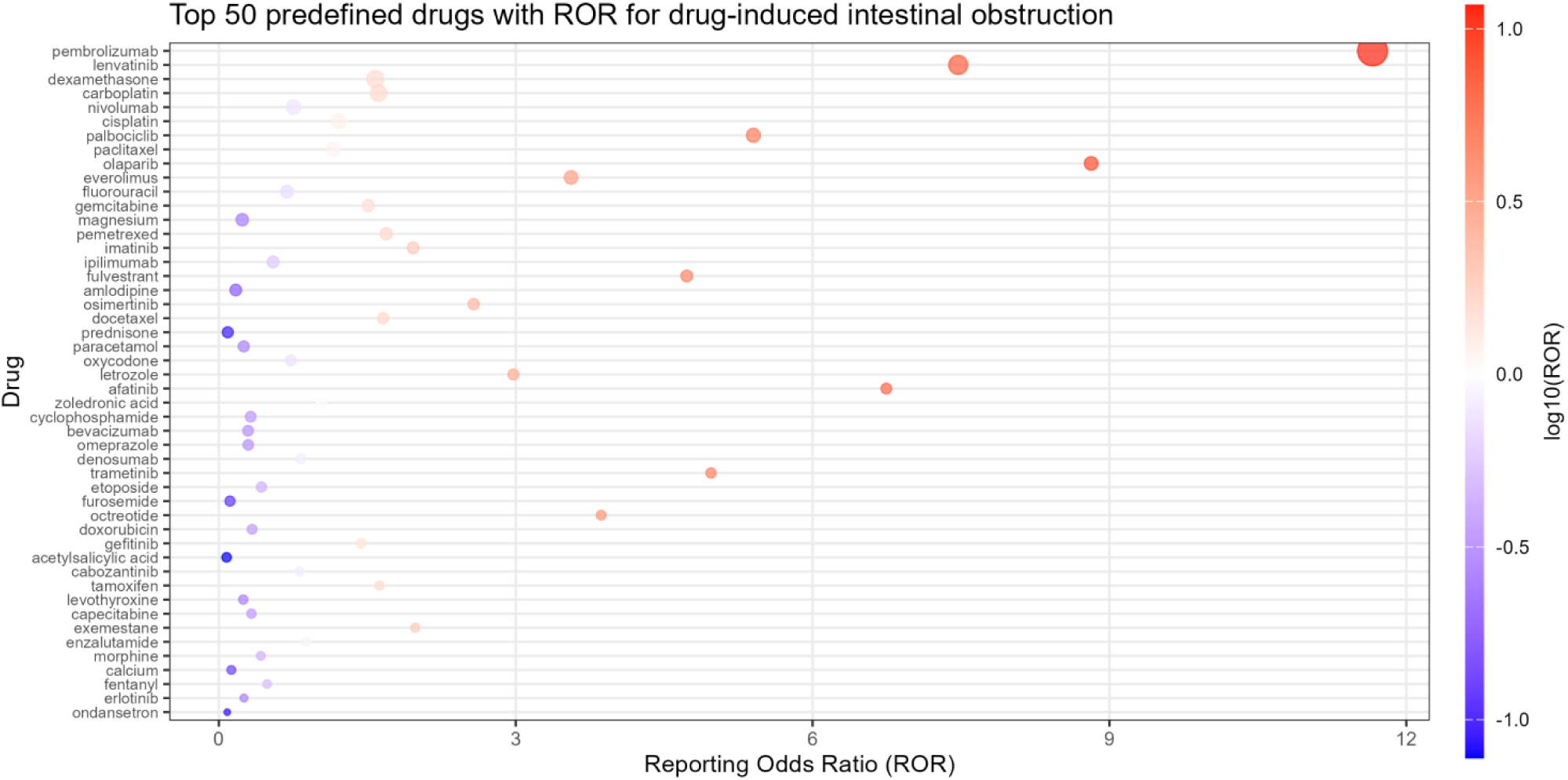
The FAERS database corresponds to the distribution of ROR signal intensity associated with malignant tumor progression in the JADER database for drugs.

## 4 Discussion

In the present study, we used FAERS and JADER to screen for drug–event signals related to malignant neoplasm progression, a rare but clinically important endpoint. Reports of hyperprogressive disease have increased over the past two decades. Does this rise mainly reflect greater clinical attention, or does it point to atypical acceleration under certain treatments? With this question in mind, we identified 50 prioritized signal drugs from large-scale spontaneous reports and summarized the clinical features of the corresponding cases. The same drug list was then re-examined in JADER to assess reproducibility across reporting systems. Importantly, cross-database consistency does not establish causality. Rather, it suggests that a “suspicious pattern” can be repeatedly captured, so our findings should be treated as hypothesis-generating safety alerts rather than precise effect estimates.

From a therapeutic spectrum perspective, antineoplastic and immunomodulating agents comprised the largest share of the 50 drugs. Many immune checkpoint inhibitors, particularly PD-1/PD-L1 antibodies, showed disproportionate reporting for tumor progression, aligning with published observations of hyperprogressive disease^[26–28]^. Nivolumab and pembrolizumab were among the drug–event pairs that warrant priority attention. Yet a key question remains: if immunotherapy can provide durable benefit for some patients, why does rapid worsening occur in others? This contrast suggests that the risk boundary may not be uniform across patients^[7][29]^.Since hyperprogressive disease was described by Champiat et al. in 2017(30) [30], retrospective studies have reported rates of roughly 5%–15% among patients receiving immunotherapy^[31][32]^. Ferrara et al. observed early accelerated tumor growth in about 16% of patients with advanced lung cancer treated with PD-1 inhibitors, with poorer early survival than chemotherapy controls ^[31]^. In hepatocellular carcinoma, Kim et al. reported an hyperprogressive disease rate of about 10% and suggested elevated LDH and liver metastases as potential risk factors^[8][26]^. Consistent with these reports, PD-1/PD-L1 inhibitors in our analysis also showed disproportionate tumor progression signals^[33][34]^. Whether similar risks extend to CTLA-4 inhibitors (e.g., ipilimumab) and newer bispecific immunotherapies remains under active debate, and the boundary of evidence is not yet fully defined; continued monitoring is therefore warranted^[35]^.

Here disproportionate progression reporting was not limited to immunotherapy. Signals were also observed for some targeted agents and conventional chemotherapies. Why should targeted therapy be part of this discussion? For first-generation EGFR tyrosine kinase inhibitors, increased reporting may occur in settings where rapid worsening follows acquired resistance^[36][37]^. Compared with hyperprogressive disease, the immune-based explanation is less defined; notably, however, rapid clinical deterioration after resistance is a familiar pattern in practice.Signals were also captured for anthracyclines. Could this mainly reflect heavier use in refractory patients with aggressive tumor biology rather than a direct drug effect? In spontaneous reports, confounding by indication and disease severity is difficult to remove. Studies comparing doxorubicin and liposomal doxorubicin have noted differences in the timing of progression across formulations^[38][39]^. Emerging modalities such as CAR T-cell therapy and cancer vaccines may also warrant vigilance, although report counts were limited in our dataset^[40–42]^. Because these treatments act on the immune system, atypical acceleration is biologically plausible. Mechanistic work has linked hyperprogressive disease to baseline immune features, including a higher proportion of senescent CD4+ T cells, and to host genetic factors such as Fcγ receptor polymorphisms^[43–45]^. Taken together, these observations suggest that rare but severe rapid worsening can occur under some novel therapies and should be recognized early in clinical practice^[31][32]^.

It is also important to clarify the endpoint itself. “Malignant neoplasm progression” is not a traditional adverse drug reaction. Increased reporting does not imply direct toxicity and may reflect lack of benefit or the natural course of cancer. Therefore, the signals identified here should be interpreted as safety signals that warrant further evaluation, not as proof of causality.A key strength of pharmacovigilance mining is its scale and broad coverage, which can reveal concerns that may be underrepresented in clinical trials^[15]^. Post-marketing surveillance, including FAERS, has identified rare but serious immune-mediated toxicities for pembrolizumab and other agents, which then informed label updates and monitoring guidance^[46]^. But a related question follows: reports reflecting lack of efficacy can also appear in FAERS, and such patterns require confirmation in epidemiologic studies or other high-quality real-world datasets before causal inferences are made.Our analysis provides a quantitative approach to evaluate tumor progression as an unconventional endpoint and may inform benefit–risk assessment. Clinically, early and repeated assessment of tumor burden after treatment initiation is important. If progression is faster than expected, treatment interruption or switching may be considered, and reporting should be encouraged to support ongoing safety assessment^[47][48]^. Closer early monitoring is particularly important in higher-risk patients, such as older individuals and those with multiple metastases, when immunotherapy or other novel treatments are initiated^[49–51]^.

### Limitations

Of course, our experiment has its limitations. First of all, spontaneous reporting systems such as FAERS are affected by under-reporting and selection bias. Only about 5%–10% of serious events may be reported^[52]^. In the field of oncology, disease progression is often explained as poor treatment efficacy or the natural progression of the disease, so it is usually not reported as an adverse reaction of a drug, which may lead to an underestimation of the true burden.

In the second place, data quality is variable, with missing and inconsistent fields. More importantly, the FDA completed the transition from Legacy AERS to FAERS in September 2012 and introduced changes in case/version structure and data fields. Trend analyses spanning 2011–2012 may therefore be affected by discontinuities, field incompatibility, or data gaps. This structural change may also explain why no 2011 entries were captured in our extracted dataset. Reporting bias is another major concern. Notoriety bias (stimulated reporting) can increase submissions for specific drug–event pairs after regulatory alerts, high-profile studies, or media attention, which can inflate reporting during certain periods and affect signal stability and interpretation^[53][54]^. Under the constraints of spontaneous reports, we could not fully separate true risk changes from shifts in reporting behavior.

Furthermore, confounding cannot be fully addressed. Treated populations may differ systematically in tumor aggressiveness, stage, and prior therapy^[55]^. Immunotherapies are often used in advanced patients with limited options, who may have faster baseline progression^[56]^. Because key clinical details (e.g., tumor type, stage, and line of therapy) are frequently missing, residual indication confounding is likely. Methods such as active-comparator–restricted disproportionality analysis may improve comparability within the same indication and reduce prescribing bias^[57][58]^. Even so, such restrictions cannot replace well-curated cohort data that capture staging and treatment pathways.

Last but not least, disproportionality signals do not establish causality. Although cross-validation in JADER supports robustness, differences in prescribing patterns, launch timing, and reporting practices across countries may remain. These findings should therefore be treated as risk hypotheses and tested in cohort or case–control studies, or in curated real-world datasets, to confirm clinical relevance and quantify potential impact^[59]^.

## 5 Conclusion

Within the scope of this study, we used malignant neoplasm progression as a pharmacovigilance endpoint to mine FAERS and JADER and identified multiple drugs with disproportionate reporting for this outcome. The signals were not randomly distributed; instead, they clustered mainly in anticancer immunotherapies and targeted agents. Notably, disproportionality analysis does not establish causality, but it can help prioritize drug–event pairs that warrant closer monitoring. Even if such patterns are not emphasized in product labels, they may still add value for early safety surveillance.

This leads to a practical question: is it sufficient for benefit–risk assessment to focus only on conventional adverse reactions? In oncology, “disease progression” and “lack of benefit” sit in a grey zone between safety and efficacy interpretation, especially for therapies with novel mechanisms. Our findings suggest that reporting and classification frameworks could be refined so this endpoint is captured more consistently. In practice, early and repeated assessment of tumor dynamics after treatment initiation may help detect unexpectedly rapid worsening.

Future work should test these hypotheses in more comparable populations. Prospective studies and well-designed real-world analyses are needed to validate signal robustness and clarify underlying biological mechanisms. Better linkage between pharmacovigilance systems, cancer registries, genomic data, and treatment pathways may enable more precise characterization of abnormal progression patterns and support individualized risk management and safer prescribing decisions.

## Data Availability

The data underlying the results presented in the study are available from https://fis.fda.gov/extensions/FPD-QDE-FAERS/FPD-QDE-FAERS.html.

## 6. Acknowledgments

We sincerely thank patients who volunteered to take part in this study.

